# Measuring Reliability in Locally-deployed Language Model Dysarthric Speech Assessments

**DOI:** 10.1101/2025.10.25.25338793

**Authors:** Ondrej Klempir, Juliana Grand Mullerova, Ales Tichopad, Radim Krupicka

## Abstract

Speech is a rich and non-invasive source of clinical information, potentially providing digital biomarkers for neurological disorders such as Parkinson’s disease (PD). Impaired articulation and reduced intelligibility are among the most pervasive PD symptoms, which has motivated research into automated, objective quantification of speech deficits. This study investigated whether metrics derived from automatic speech recognition (ASR) and large language models (LLMs) can quantify speech intelligibility and describe clinical severity. Recordings of fixed read text from patients with PD and healthy controls (HC) were transcribed and evaluated using conventional ASR error measures (such as Word and Character Error Rate), a proposed Mistral-based LLM intelligibility score, and BERT-derived typo metrics (BERT, short for Bidirectional Encoder Representations from Transformers). Group-level discriminability between PD (N = 16) and HC (N = 21) was low (Mann–Whitney p > 0.05; Random Forest obtained Receiver Operating Characteristic Area Under the Curve of 0.66; leave-one-subject-out evaluation), indicating that transcript-level features alone offer limited classification abilities. Importantly, the LLM-derived intelligibility score demonstrated excellent repeatability across five runs (intra-class correlation; ICC = 0.97, Cronbach’s α = 0.98) and showed strong correlations with ASR error metrics. Both LLM and ASR measures significantly correlated (p < 0.05; Spearman’s rank correlation) with common rating clinical scales (Hoehn and Yahr scale and Unified Parkinson’s Disease Rating Scale), whereas BERT-typo parameters did not. These findings suggest the use of LLMs as tools for generating reference-free intelligibility scores that reflect disease severity.

## 1. Introduction

Artificial Intelligence (AI) is transforming how we learn, design, and deploy solutions. Recent AI scoring systems now integrate automated rationale generation, moving beyond numerical outputs to include interpretable explanations that enhance transparency and trust [1]. Powered by large language models (LLMs), these systems generate both scores and reasoning, creating self-explainable frameworks that improve understanding and application [2, 3, 4].

In medicine, AI severity scores offer clinicians quantitative estimates of pathology likelihood. Yet, limited attention has been paid to how these scores are produced or interpreted, raising concerns about reliability and transparency [5]. Six main limitations have recently been identified [5]: (1) variability across systems, (2) variability within systems, (3) variability between experts, (4) variability within experts, (5) unknown score distributions, and (6) perceptual challenges. Consistency between human and AI evaluators should be assessed using inter-rater reliability or Bland–Altman analysis [6]. Encouraging progress has been shown in clinical risk computation using LLMs with validated risk scores [7], though intra-model repeatability is under assessed. Moreover, a recent prospective clinical study compared multiple LLMs in orthopaedic diagnostics and found that GPT-4o achieved the highest diagnostic sensitivity (92.3%), significantly outperforming other models such as Gemini and Llama-3.1 [8]. Inter-model agreement was only moderate among GPT-4 versions and was relatively low across other models, emphasizing the need for broader benchmarking and inclusion of additional systems such as Microsoft Copilot in future research.

A rapidly advancing domain in current biomedical research is voice phenotyping and vocal biomarker research, where speech and voice signals are analysed to detect and monitor health conditions using AI and speech foundational models [9, 10, 11]. Advances in signal processing and machine learning now enable the extraction of biomarkers for early detection and monitoring of neurodegenerative disorders, particularly in digital phenotyping of Parkinson’s disease (PD) [12]. Impairment of speech production and intelligibility is a common symptom in PD [13], and recent findings emphasize that language plays a critical role in early PD detection from speech [14]. Speech analysis offers a promising alternative to traditional data sources. Unlike administrative or claims data, it can reveal subtle motor and cognitive changes long before clinical diagnosis, providing a potentially superior early signal for understanding and optimizing PD pathways.

Our previous study investigated automated speech recognition (ASR) in PD, evaluating speech intelligibility and severity using the speech foundation model Wav2Vec 2.0-base, non-fine-tuned [15]. ASR enables the transcription of speech into text, which can subsequently be analysed by (large) language models. Crawford [16] employed Whisper for ASR and demonstrated that PD can be automatically detected from spontaneous speech transcripts, without using acoustic features, with up to 78% accuracy when distinguishing PD patients from healthy controls (HC). In addition to classification, their regression model predicted Movement Disorder Society–Unified Parkinson’s Disease Rating Scale Part III (MDS-UPDRS-III; UPDRS-III) scores from transcript embeddings.

Fine-tuning foundation speech models on dysarthric speech remains challenging due to overfitting from limited dataset sizes. A recent study proposed a multitask learning framework to mitigate overfitting during fine-tuning [17], improving both in-corpus and cross-corpus detection accuracy. Similarly, a two-stage data augmentation approach has been shown to enhance ASR performance for dysarthric speech [18], achieving improved Word Error Rate (WER), i.e. one of the key metrics for ASR evaluation, across different intelligibility levels.

In addition to traditional minimum-edit distance metrics such as WER, which require reference transcriptions for comparison, recent studies have begun to link clinical variables in PD directly to ASR and possibly language model outputs. Castelli et al. [19] used LLMs to predict the Neuropsychiatric Fluctuations Scale (NFS) in both ON and OFF medication states, based on spontaneous speech during monologue tasks. Tröger et al. [20] introduced the SB-M intelligibility score, an automatic proxy for speech intelligibility that correlates with clinical anchor measures such as the UPDRS. Furthermore, [21] highlighted limitations of traditional ASR metrics for dysarthric and dysphonic speech, noting that WER may not reflect perceived intelligibility when semantic alignment outweighs word-level accuracy. To address this, a new evaluation metric combining Natural Language Inference (NLI), semantic similarity, and phonetic similarity was proposed to better capture speech intelligibility.

Herein, we propose an LLM-based pipeline implementing Mistral-7B to assess PD speech using a single score, without requiring reference text. We evaluated its repeatability, reliability and validity against classical ASR metrics and a language-model-based estimator of detected typos and errors.

## 2. Methods

### 2.1. Dataset

We used the MDVR-KCL speech corpus. The set analysed comprised of 21 HC and 16 PD participants. Audio files were available as WAV recordings. Detailed acquisition procedures are described in the MDVR-KCL documentation [22]. For each PD subject, the following clinical scores were available: Hoehn & Yahr (H&Y), UPDRS Part II (item 5), and UPDRS Part III (item 18). We used both of these individually and as a composite score (sum of H&Y and the two UPDRS items). Note that H&Y is a global staging measure reflecting overall disease progression (early vs. advanced PD; and often correlates with disease duration) and should not be interpreted as a direct indicator of speech impairment. In contrast, UPDRS includes targeted motor and functional items that captures deficits more directly relevant to speech and intelligibility, and is therefore a more appropriate clinical indicator for speech-focused analyses.

For consistency across subjects we selected the read-text task only. For each recording, we manually trimmed and retained only the canonical passage The North Wind and the Sun (from “The North Wind and the Sun were disputing which was the stronger…” to “…to confess that the Sun was the stronger of the two.”), for which a gold standard reference exists.

### 2.2. Speech-to-Text (ASR)

The ASR task was to automatically transcribe the speech into text. Transcript quality is critical because downstream analyses operated on ASR outputs. We used the open-source Wav2Vec 2.0 base model [23] from Hugging Face (facebook/wav2vec2-base-960h). This model provides self-supervised speech representations and has a strong baseline performance for English ASR. We used the raw English 960-hour model (not fine-tuned for multilingual or dysarthric speech). Audio input was resampled to 16 kHz prior to feeding into the model. All recordings were processed offline on a computer with i5-7400 3GHz CPU and 16GB RAM.

### 2.3. Minimum-edit distance metrics

We evaluated ASR output with deterministic minimum-edit distance metrics using the JiWER Python library [24]. These measures required a reference transcription and included: Word Error Rate (WER), Match Error Rate (MER), Word Information Preserved (WIP), Character Error Rate (CER), Word Information Lost (WIL). We considered WIL as particularly relevant for clinical/dysarthric speech, as it serves as a more sophisticated alternative to WER that reduces sensitivity to small, non-meaningful errors (e.g. punctuation or single-letter differences) [25].

All metrics were computed on the aligned reference–hypothesis pairs using standard minimum-edit distance algorithms implemented by JiWER.

### 2.4. Typo detection: DistilBERT

We used a BERT-family (BERT, short for Bidirectional Encoder Representations from Transformers) encoder model to detect misspellings/typos in ASR transcripts. BERT is an encoder-only transformer that produces contextual embeddings and can be fine-tuned for token-level classification. We employed distilbert-base-multi-cased-finetuned-typo-detection (with its corresponding tokenizer), which was fine-tuned on the GitHub Typo Corpus (a large multilingual dataset of misspellings and corrections [26]). The model returns token-level labels (e.g., typo or ok) and associated confidence scores. For each transcript, we computed three metrics:

- Percent typos — percentage of tokens labelled as typos.
- Absolute number of typos — raw count of tokens labelled as typos.
- Score variability — standard deviation of the model’s score field across tokens, used as a proxy for confidence variability.

Token classification inference is deterministic unless model randomness is explicitly enabled. Thus, typo detection was treated as a deterministic baseline for downstream comparisons.

### 2.5. LLM-based scoring (Mistral-7B, local)

We implemented a local LLM-based Python pipeline that assigns a single intelligibility/quality score (0 worst – 10 best) to each transcript without requiring a reference text. A local Ollama server (Ollama v0.11.11) hosted the model and was accessed via LangChain [27]. We used Mistral-7B (4.4 GB) as the decoder-only generative LLM. Mistral operates as a causal (decoder) transformer similar in family to LLaMA/GPT.

The scoring prompt used for each transcript was:

*“You are an expert on evaluating Parkinson’s disease speech, with access to the following transcription resulting from Automated Speech Recognition (ASR)*.

*Please evaluate the input text (lexical and syntactic, grammar, composition, intelligibility etc*.*) and suggest a score - one number on a scale from 0 (the worst) to 10 (the best) - based ONLY on the information in the transcriptions. Output will be a string with one numeric value*.

*INPUT: {transcription}”*

The model response was parsed to extract the numeric score. No external reference transcription was provided to the LLM. Because decoder-only models can produce variability across independent runs even with identical prompts, we executed multiple independent runs per transcript (n = 5) to characterise intra-model variability and reliability.

All LLM computations were executed locally on a MacBook Air equipped with an Apple M1 processor and 8 GB RAM, using CPU-only inference (no GPU acceleration was required).

### 2.6. Statistical analysis and learning

All statistical analyses were conducted in Python. A significance threshold of p < 0.05 was used.

#### 2.6.1. Reliability and agreement

We assessed the consistency and agreement of the Mistral-derived scores by computing intraclass correlation coefficient (ICC(3,k)) using the pingouin package [28]. Interpretation of ICC followed commonly used thresholds (<0.40 poor, 0.40–0.59 fair, 0.60–0.74 good, 0.75–1.00 excellent) as in prior psychometric work [29]. To complement the ICC analysis we computed Cronbach’s alpha to quantify internal consistency across repeated LLM runs, and performed Bland–Altman analyses (including plots and limits of agreement) between LLM replicates to visualise systematic bias and dispersion across repeats.

#### 2.6.2. Correlation analyses

Monotonic associations between ASR-derived metrics and clinical scores were evaluated using Spearman’s rank correlation. Specifically, we tested the correlations between established ASR error measures (WER, WIL, CER etc.), typo-detection metrics (percent and absolute typos, score variability), and the single-number LLM intelligibility score against clinical anchors (H&Y, UPDRS items and the composite clinical score). In addition, Spearman correlations were used to compare the different ASR assessment approaches with one another (for example, WER versus WIL, and WIL versus the Mistral score) to characterise convergent and divergent validity among the metrics.

#### 2.6.3. Group differences and discriminability

Group differences between HC and PD participants for individual ASR-derived metrics were tested with the nonparametric Mann–Whitney U test [30]. Furthermore, to quantify discriminative performance, we trained a Random Forest classifier (RF; n_estimators=200) [31] and evaluated it with Leave-One-Out cross-validation (LOOCV). Reported performance measures included Receiver Operating Characteristic Curve, Area Under the Curve (ROC-AUC), balanced accuracy, accuracy, and average precision (PR-AUC). Classifiers were trained on combined feature sets to evaluate its information content and discriminative value.

## 3. Results

### 3.1. Low discriminability between HC vs. PD

At the transcript level, none of the evaluated ASR- or LLM-derived features showed statistically significant differences between HC and PD participants (Fig. 1). When combining all nine features in a RF classifier, only moderate discrimination was achieved (ROC–AUC = 0.66, balanced accuracy = 0.62, accuracy = 0.62, average precision = 0.71).

**Fig. 1.**
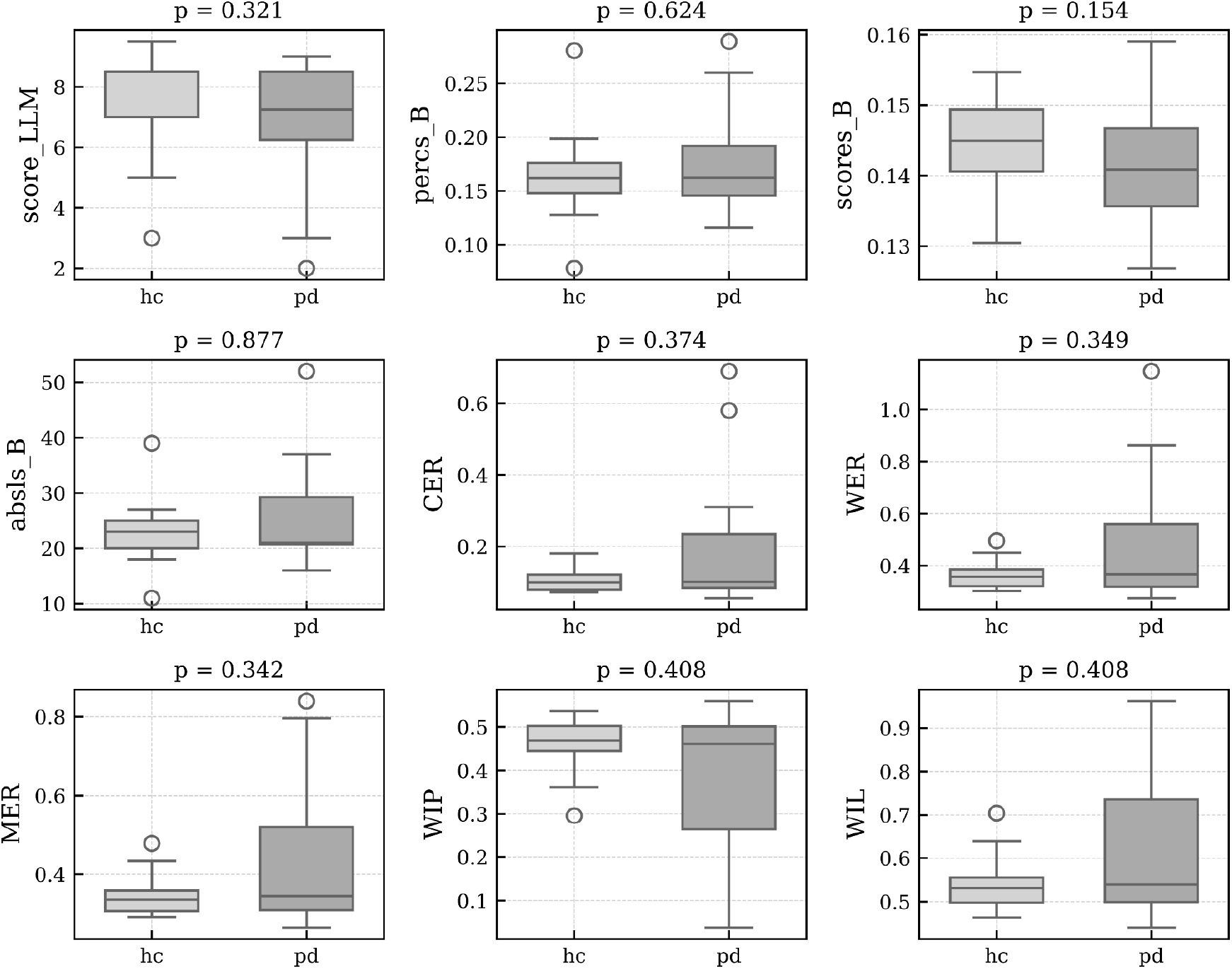
Boxplots comparing HC and PD groups across nine transcript-derived metrics. Panels include minimum-edit distance measures (CER, MER, WER, WIL, WIP), the LLM-based intelligibility score (score_LLM), and typo-detection metrics derived from the BERT model (percs_B, absls_B, scores_B). Reported p-values correspond to Mann–Whitney U tests. No feature reached statistical significance (all p > 0.05).

### 3.2. Excellent repeatability of the LLM intelligibility score

The LLM-derived intelligibility score verified excellent repeatability across five independent runs (ICC(3,k) = 0.97, 95% CI [0.96–0.99], p < 0.001). Cronbach’s α = 0.98 confirmed strong internal reliability.

Fig. 2A shows boxplots of scores across five repetitions with no apparent distributional shift. In addition, Fig. 2B presents a correlation heatmap, where pairwise Spearman coefficients ranged from ρ = 0.85 (Rep 3 vs Rep 5) to ρ = 0.92, confirming high inter-run consistency. The Bland–Altman plot (Fig. 2C), comparing Rep 1 and Rep 2, yielded a mean bias of -0.14, with all but one observation within the 95% limits of agreement.

**Fig. 2.**
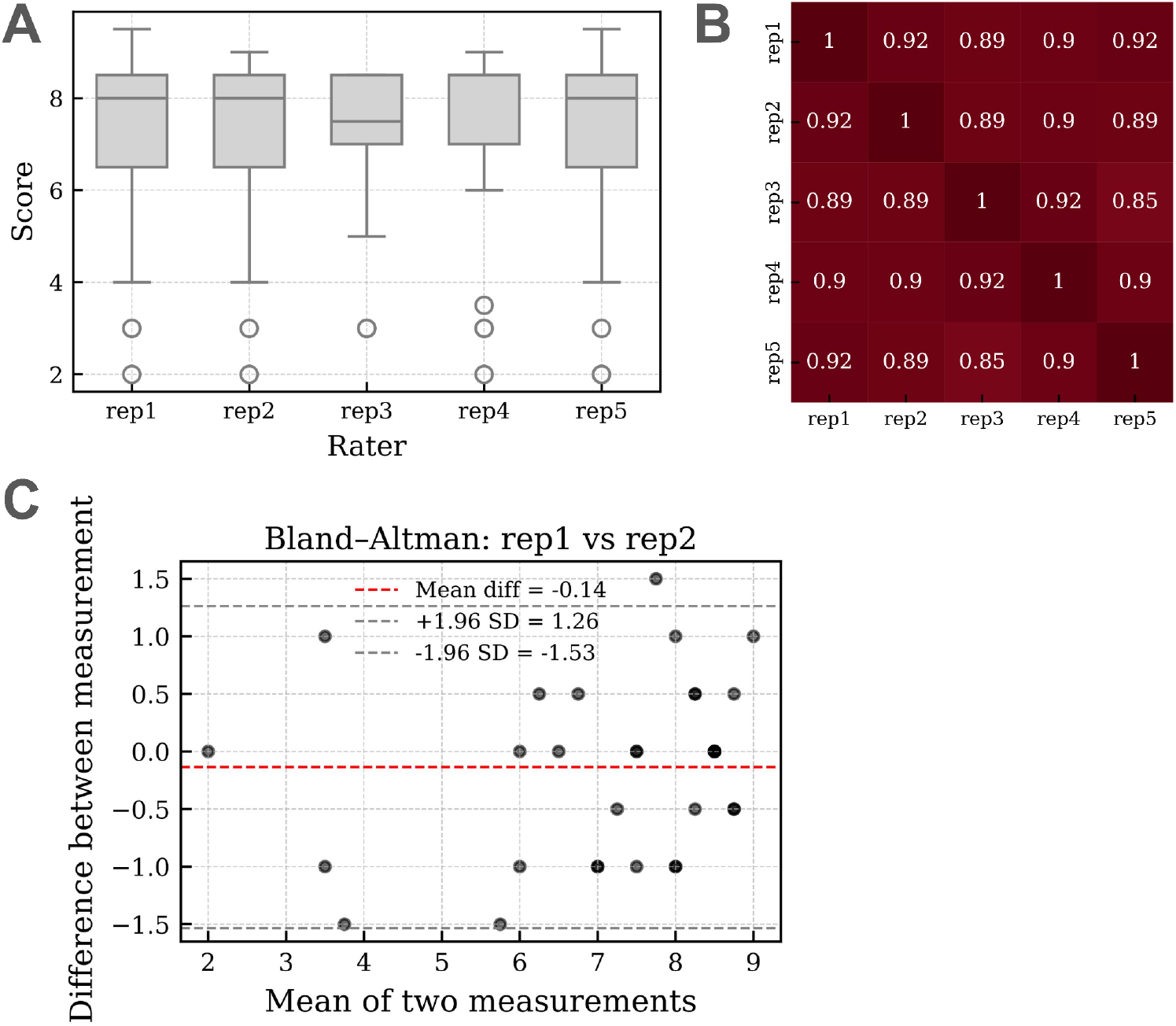
Repeatability analysis of the LLM-based intelligibility score. (A) Boxplots showing score distributions across five repetitions. (B) Spearman correlation heatmap showed high inter-run agreement (ρ ≥ 0.85). (C) Bland–Altman plot comparing Rep 1 and Rep 2. Overlapping points indicate multiple subjects with identical values.

For repetition #1, the mean runtime across 37 samples was 9.85 ± 5.46 seconds (min. 1.73 seconds, max. 21.53 seconds). The first run was slower due to model initialization, after which runtimes stabilized below 15 seconds.

### 3.3. Inter-metric correlations for intelligibility parameters

Overall, a strong internal agreement was observed among intelligibility-related feature sets (Fig. 3). As expected, features within the same group (e.g., minimum-edit distance measures such as MER, CER, and WER) showed the highest inter-correlations. Importantly, the LLM-based intelligibility score correlated significantly with all minimum-edit distance metrics and with the BERT-derived score_B parameter. No significant associations were found with the absolute or percentage counts of typos in transcripts.

**Fig. 3.**
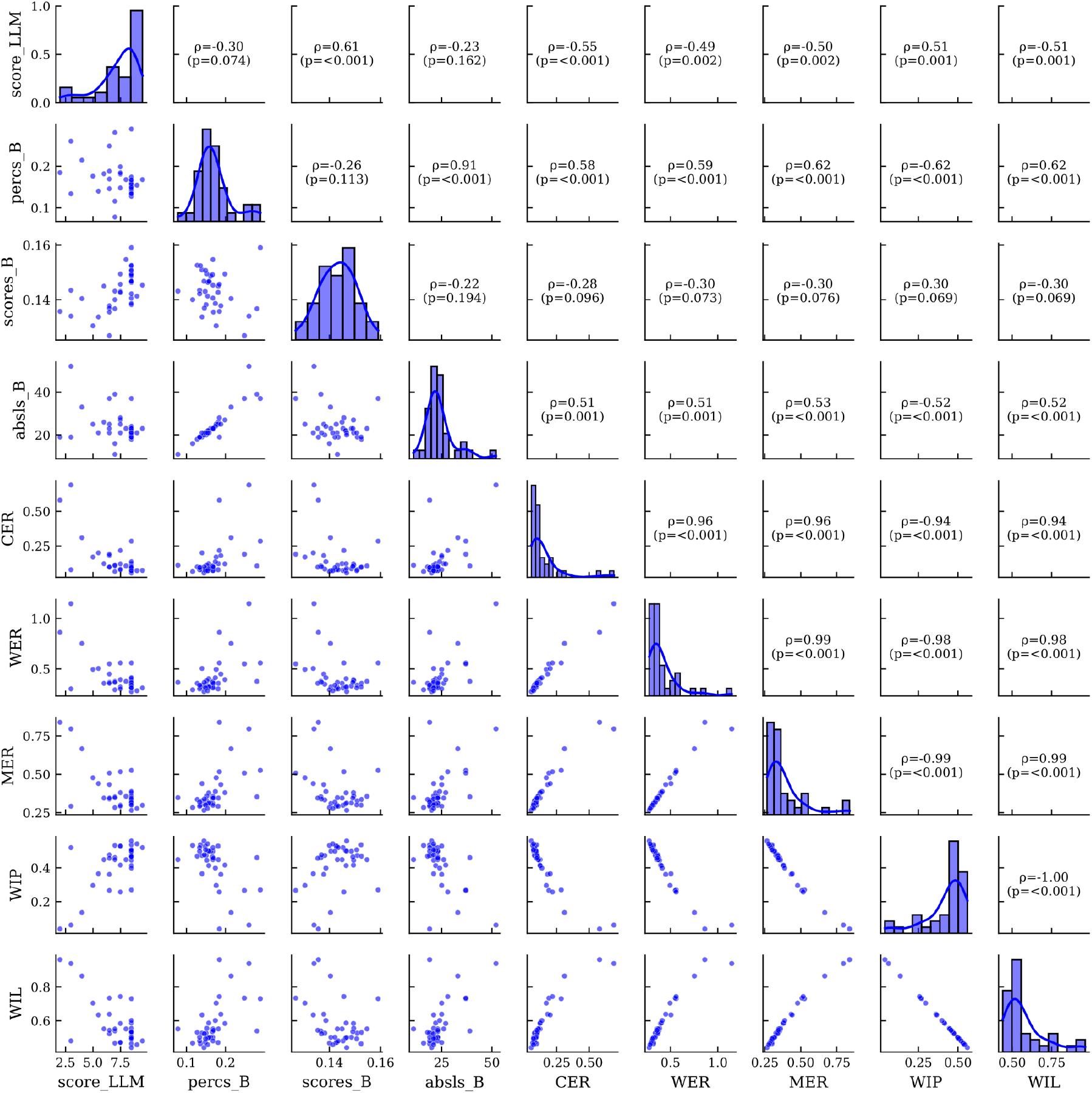
Correlation matrix of intelligibility-related feature sets.

### 3.4. Associations between ASR and clinical scores

All minimum-edit distance measures and the LLM-based intelligibility score showed significant correlations with PD severity indicators, including UPDRS-II, UPDRS-III, H&Y, and their composite score. In contrast, none of the BERT-derived typo metrics correlated with any clinical variable. Selected associations are illustrated in Fig. 4.

**Fig. 4.**
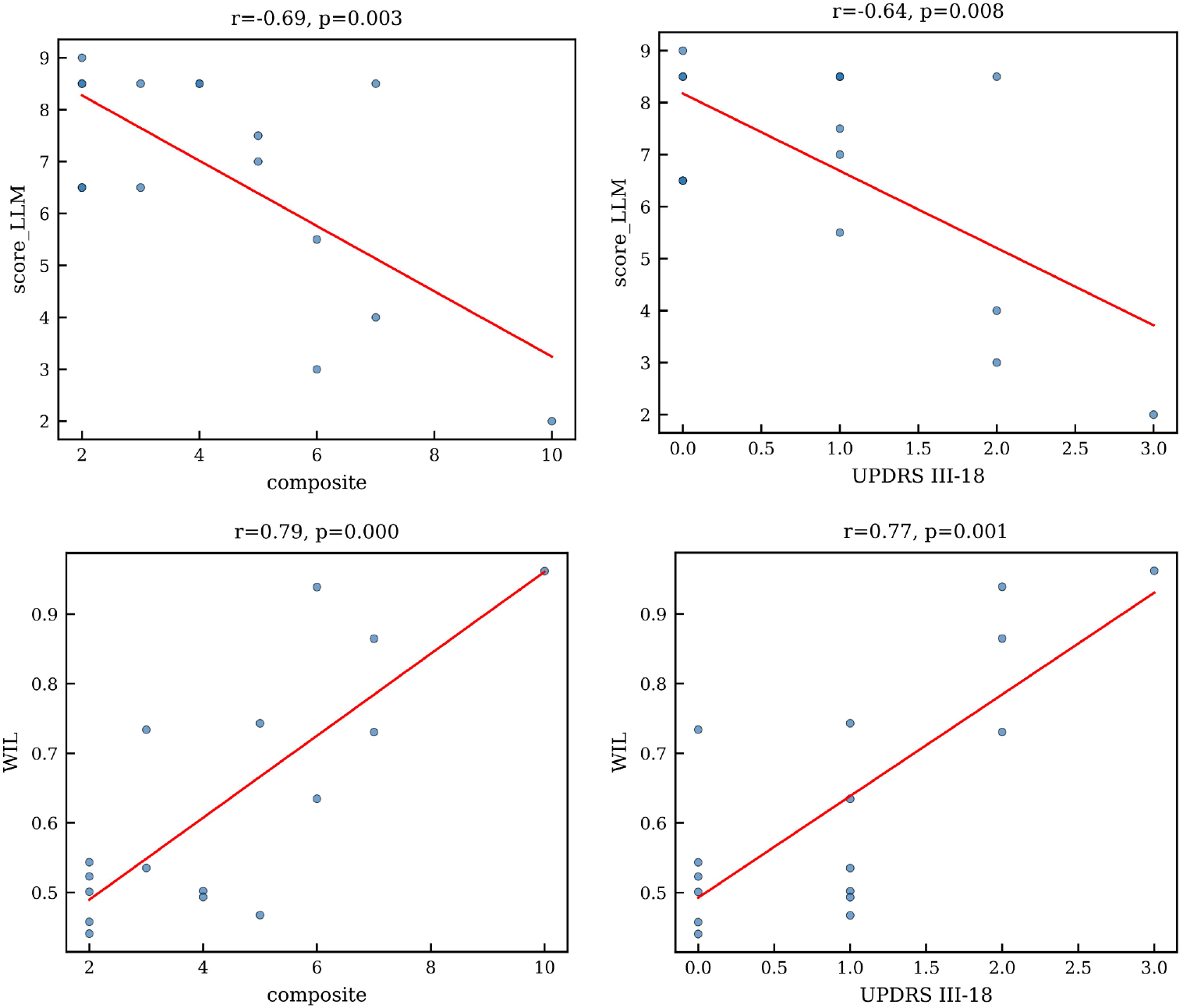
Associations between intelligibility metrics and clinical severity scores. Scatter plots show relationships between (top left) LLM-based score and composite clinical score, (top right) LLM-based score and UPDRS-III item 18, (bottom left) WIL and composite score, and (bottom right) WIL and UPDRS-III item 18. For LLM-based scores (top panel), overlapping points indicate multiple subjects with identical scores.

## 4. Discussion

In this study, we developed a solution that integrates ASR using Wav2Vec 2.0, transcript-level distance metrics features, a DistilBERT-based typo detection, and scoring from a local Mistral-7B language model, to quantify speech intelligibility and describe clinical severity. While these feature sets did not reliably discriminate PD from HC in the read-text task, they did relate robustly to clinical severity measures. The findings suggest that both ASR-derived and LLM-based metrics capture clinically relevant aspects of speech impairment, whereas typo frequency alone does not reflect disease severity.

The lack of statistically significant group separation (HC vs. PD) at the transcript level likely reflects several factors. First, our task used a constrained read passage (The North Wind and the Sun), which reduces inter-subject linguistic variability compared with spontaneous monologues. Crawford [16] demonstrated high classification accuracy from transcripts (Spanish PC-GITA dataset) when using spontaneous speech, which is content-rich and participant-specific. In those less controlled settings, HC might typically produce richer and more variable utterances, which may increase between-group differences in the linguistic feature space. Second, as shown in Fig. 1, many HC samples exhibited low WER/CER and some PD samples also had lower WER/CER, therefore ASR measures failed to capture deeper, subtler speech impairments that do not manifest as word substitutions or deletions. The performance of any downstream model is therefore highly dependent on the quality of the underlying transcription. Although the Wav2Vec 2.0–based transcriptions used in this study were generally reliable, alternative ASR systems such as Whisper could be explored in future work. Third, the sample size presented might limit statistical power and the ability to detect small group effects.

The LLM-derived intelligibility score showed excellent repeatability (ICC and Cronbach’s α) and converged with multiple ASR error metrics, including WIL, a metric designed to emphasize meaningful word-level information. These convergent associations and the significant correlations with UPDRS and H&Y indicate that LLMs can extract features from transcript text that track clinical severity. In contrast, the BERT-based typo-detection metrics (absolute and percentage typos) did not correlate with clinical scores. This may reflect the nature of the training corpus for the typo model (large, noisy GitHub-derived edits) and suggests that simple misspelling counts are insufficient as proxies for intelligibility or motor impairment.

To further validate repeatability, the presence of a single sample outside the Bland–Altman limits (±1.96 SD) among 37 subjects (Fig. 2C) indicates high overall repeatability, suggesting that only one measurement showed minor deviation beyond expected random variation. The outlier corresponded to a HC subject who obtained a score of 8.5 in repetition #1 and 7.0 in repetition #2, representing a difference of 1.5 points (approximately 15% change on the 0–10 scale). Such deviation likely reflects normal intra-individual variability.

Our results therefore point to a realistic and clinically relevant use-case. While transcript-only features may be insufficient for binary screening of PD in constrained read tasks, LLMs can provide reference-free proxy scores that quantify severity-related linguistic degradation. This is attractive for text-based monitoring (e.g. telehealth, phone screening) where raw audio processing may be impractical.

Recent research has examined agreement across closed-source LLMs. Ming et al. [32] evaluated inter-model agreement among OpenAI’s o1 and o3, and DeepSeek-R1, quantified using Cohen’s kappa. They reported moderate–high agreement between R1 and o3, but only slight agreement with o1 in the context of ophthalmic diagnosis and reasoning. In comparison, ASR–based evaluation presented herein likely represents a simpler and more repeatable task. A further strength of the present work is that it demonstrated reliable performance even with a compact, locally deployed model (Mistral-7B), implying that larger LLMs may perform equally well or better on similar linguistic evaluation tasks. Atil et al. [33] highlighted that model reliability and calibration across reasoning scales are essential for ensuring consistent decision behavior in LLMs, reinforcing the importance of agreement analysis in this domain.

Some possible limitations should be indicated. While the PD subgroup remains relatively small, the overall sample of 37 participants can be considered robust, in the context of having carefully curated ground-truth transcriptions. We relied on the Wav2Vec 2.0 model which was not fine-tuned for dysarthric speech, and in general, it likely limited transcription accuracy for severely affected speakers. In our opinion, the use of Wav2Vec 2.0 without fine-tuning can alternatively be viewed as a methodological feature rather than a drawback. While a fine-tuned model could potentially improve transcription accuracy for severely affected speakers, it might also normalize or “correct” speech errors, i.e. possibly masking clinically relevant markers of disease severity. This suggests that, for tasks aiming to quantify or monitor speech impairments, general-purpose foundational ASR models may be preferable. Furthermore, although we ran the LLM scoring multiple times to quantify repeatability, the broader distributional properties of LLM-derived scores (per-subject and per-group) require more extensive repetition and calibration. Finally, our pipeline used only transcript-derived features and omitted acoustic features (such as prosody or spectral measures), removing a rich source of diagnostic information known to perform well on MDVR dataset [34].

For future research, comparative benchmarking across multiple LLMs (including proprietary models or systems like Microsoft Copilot/GPT-4o/GPT5.0) would clarify model-specific advantages. Additional methodological work should investigate the distribution and calibration of LLM scores (more repeated runs, uncertainty estimation, and ensembling), and examine longitudinal sensitivity to disease progression and medication state (ON/OFF).

## 5. Conclusion

We designed and evaluated an LLM-based, reference-free intelligibility score derived from ASR transcripts in PD. Although transcript-level features showed limited ability to discriminate PD from HC, the LLM score reliably tracked clinical severity and was highly repeatable across independent runs (high ICC and Cronbach’s α). The LLM score converged with established ASR error metrics (e.g. WER, WIL, CER) and explained severity within the PD subgroup, demonstrating that intelligibility can be assessed from text without a reference transcript. The proposed LLM-derived score therefore constitutes a novel clinical biomarker for speech impairment in PD and a practical building block for text-based monitoring and telehealth applications.

## Data Availability

All data produced in the present study are available upon reasonable request to the authors

## Acknowledgement

This study was supported by the project of the National Institute for Neurological Research (Programme EXCELES, ID Project No. LX22NPO5107), funded by the European Union, Next Generation EU. Part of this research (primarily the LLM experiments) was supported by project CZ.02.01.01/00/23_025/0008743, funded by the European Union under the Operational Programme Johannes Amos Comenius (OP JAK).

## Notes

### Competing Interest Statement

The authors have declared no competing interest.

### Author Declarations

The study used ONLY openly available human data. The dataset used is available at the following URL: https://zenodo.org/records/2867216.

